# The association between nighttime fasting intervals and depression from early pregnancy to postpartum

**DOI:** 10.64898/2026.07.24.26358661

**Authors:** Takashi Yoshimasu, Christina M. Personette, Namhyun Kim, Rya Clifton, David Phan, Mehak Behal, Riley J. Jouppi, Andrea B. Goldschmidt, Christine C. Call, Lydia B. Brown, Shruti Kinkel-Ram, Rachel P. Kolko Conlon, Michele D. Levine, Marquis Hawkins

## Abstract

**Purpose:** Nighttime fasting intervals (NFIs; i.e., the longest period between the last eating episode of the day and the first of the next day) are associated with depressive symptoms in non-pregnant populations. However, this chrono-nutrition indicator has rarely been explored in the perinatal period, when individuals experience dramatic physiological and psychological changes. We explored potential associations between NFIs and depressive symptoms during the perinatal period.

**Methods:** This secondary analysis included 232 individuals with pre-pregnancy body mass index (BMI) ≥25 kg/m². Participants were assessed during early-pregnancy, mid-pregnancy, and at 6-months postpartum. NFIs and breakfast time were estimated from 24-hour dietary recall interviews, and depressive symptoms were self-reported using the Center for Epidemiologic Studies Depression Scale. NFIs were categorized into three classifications to represent short, middle, and long NFIs. Longitudinal associations were evaluated using mixed effects linear models with interaction terms between NFIs and each timepoint.

**Results:** NFIs remained largely stable at the group level across the three study visits (*M* [*SD*] = 12.7 [2.75] hours). Although NFIs were not significantly associated with depressive symptoms, long NFIs showed geometric mean ratio of 0.84 (95% CI: 0.64–1.11) relative to middle NFIs at mid-pregnancy.

**Conclusions:** NFIs and depression were not associated among this perinatal sample. Future studies should explore circadian eating patterns (e.g., the timing of the last eating episode relative to dim-light melatonin onset), different timepoints (e.g., late pregnancy), and pregnant individuals across the weight spectrum.

**Article Highlights:**

1. Nighttime fasting intervals are associated with depressive symptoms in non-pregnant populations, yet their role during the perinatal period remains unclear. We investigated NFIs and depressive symptoms across early-pregnancy, mid-pregnancy, and 6 months postpartum in women with overweight or obesity. NFIs were largely stable across visits, while depressive symptoms, measured by the Center for Epidemiologic Studies Depression Scale, improved over time.
2. NFIs were not significantly associated with depressive symptoms during the perinatal period overall. However, in mid-pregnancy, shorter fasting intervals were associated with approximately 10 percents higher predicted probability of depression compared to longer intervals, suggesting a potentially clinically meaningful difference that warrants further investigation.
3. After adjusting for sleep quality, longer NFIs were significantly associated with lower depressive symptoms in mid-pregnancy, which highlights sleep quality as an important covariate. Future studies with larger samples and objective measures of chronotype, sleep, and depression are needed to clarify the role of NFIs in perinatal mental health.

## Introduction

Depression during the perinatal period (i.e., pregnancy through the first year postpartum) is highly prevalent, with major depression affecting ∼6.1% and minor depression affecting ∼16.6% U.S. individuals during pregnancy (Ashley *et al*., 2016) and depressive symptoms affecting ∼19% of U.S. during the postpartum period (Wang *et al*., 2021). Depression and depressive symptoms during pregnancy are both associated with preterm birth, gestational diabetes, and preeclampsia (Stewart, 2011). Postpartum depression is further associated with elevated risks for maternal suicidality, breastfeeding difficulties, and impaired mother-infant bonding (Stewart and Vigod, 2016). Perinatal depression is multifactorial; with risk factors spanning social determinants, health-related conditions, and health behaviors (Yin *et al*., 2021; Liu, Wang and Wang, 2022). Notably, modifiable health behaviors, including poor diet, poor sleep, or smoking during pregnancy, have been shown to explain approximately 32% of the variance in depressive symptoms during pregnancy (Van Lee *et al*., 2020), suggesting that health behaviors may be potential targets for intervention.

Recent research has suggested that intermittent fasting (IF), a dietary approach that regulates the timing and interval of food intake without altering caloric consumption, may improve mental well-being through changes in neural signaling and molecular pathways (Igwe *et al*., 2021). Time-restricted eating (TRE), a form of IF restricting the time-window for energy intake, has gained particular attention in this context. For example, a TRE regimen after a set time for 50 days was associated with improvements in depression scores in healthy adults (Huo *et al*., 2025). Similarly, prolonged fasting has been associated with improvements in cognitive-affective symptoms in adult patients with major depressive disorders (Stapel *et al*., 2022). Together, these findings suggest longer fasting intervals, without necessarily altering total daily calorie consumptions, may improve depressive symptoms. However, the fasting protocols used in these studies often involved fasting intervals exceeding 18 hours and intervention periods lasting more than one month, which limits their direct applicability for perinatal populations who experience dramatic changes in energy needs, sleep, gastrointestinal symptoms, metabolic physiology, and fetal nutritional demands (Haddad-Tóvolli and Claret, 2023).

Related to TRE, nighttime fasting intervals (NFIs) may offer a more naturalistic measure of eating timing. NFIs are typically defined as the interval between the last eating episode of one day and the first eating episode of the following day. Unlike prescribed fasting interventions, NFIs reflect habitual daily eating patterns and may capture dimensions of behavioral regularity, circadian alignment, and overnight metabolic rest. Irregular or extremely long or short NFIs may suggest maladaptive health behaviors, such as skipping breakfast (Smith *et al*., 2010) or night eating after bedtime (Wang *et al*., 2024). Several studies in non-pregnant populations have investigated associations between NFIs and health outcomes, including metabolic health and depression (Parr *et al*., 2020; Palomar-Cros *et al*., 2021; Bravo-Garcia *et al*., 2024). However, findings on NFIs and depressive symptoms have been mixed. For example, in a nationally representative sample of US adults, both short and long NFIs were associated with higher risk for depression (Chen *et al*., 2025), whereas a study among airline personnel found that longer NFIs were associated with lower depressive symptoms (Zhang *et al*., 2024). These conflicting findings may reflect heterogeneity in population characteristics, dietary assessment, and the possibility that NFIs have non-linear associations with mood. Moderate overnight fasting may have protective effects by modulating the gastrointestinal system and central nervous system through the gut-brain axis or by reducing neuroinflammation (Zhang *et al*., 2024). In contrast, excessively prolonged NFIs may trigger stress responses in the brain, disrupt intestinal flora homeostasis, and negatively affect immune function (Chen *et al*., 2025). Taken together, the relationship between NFIs and depression may depend on whether longer overnight fasting reflects a stable, health-promoting eating rhythm or a marker of insufficient intake, irregular eating, or physiological burden.

Despite growing interest in fasting-related eating patterns and mood, existing studies have largely focused on nonpregnant populations, and evidence in perinatal populations remains limited. To date, a few studies have examined NFIs in pregnant populations (Loy *et al*., 2017; Skarstad *et al*., 2024), and the association between longer NFIs and lower blood glucose levels was suggested. To our knowledge, no study has investigated associations between NFIs and depressive symptoms during the postpartum period even though NFIs may represent a low-burden approach that does not require explicit caloric restriction. Therefore, the present study aims to examine the association between NFIs and depressive symptoms across the perinatal period, including postpartum.

## Materials and Methods

### Study design and setting

The present study is a secondary analysis of a prospective cohort study designed to assess psychosocial factors associated with gestational weight gain among pre-pregnancy overweight or obese perinatal individuals. Participants were recruited between September 2012 to January 2017 from obstetric clinics in Pittsburgh, PA, USA. Detailed information on the parent study can be found elsewhere (Levine *et al*., 2023).

### Participants

Pregnant people (*n* = 257) were enrolled in the study between 12 and 20 weeks of gestation. Inclusion criteria were: (1) ≥ 14 years of age, (2) pre-pregnancy body mass index (BMI) ≥ 25 kg/m^2^, and (3) singleton pregnancy. Exclusion criteria were: (1) type I diabetes, (2) use of medications and/or diagnosis of conditions known to influence weight, (3) concurrent participation in a weight management program, or (4) self-report of psychiatric symptoms requiring immediate treatment. Participants with missing NFI or depressive symptom data at more than one timepoint were additionally excluded for the purposes of the current study, resulting in a final analytic sample of 232 participants. The University of Pittsburgh’s Institutional Review Board approved all study procedures.

Participants provided written informed consent or assent before enrollment.

### Procedures

The present analysis examined variables assessed at 12-20 weeks (i.e., early-pregnancy), 27-30 weeks (i.e., mid-pregnancy), and 6-months postpartum, including 24-hour dietary recalls collected via telephone for two days and self-report measures of psychiatric symptoms. The Pittsburgh Sleep Quality Index (PSQI; Buysse *et al*., 1989) was added later in data collection.

### Variables

#### Nighttime fasting interval

We defined NFI as the longest interval between the last eating episode of one day and the first of the next, based on participants’ self-reported identification of their first and last eating episodes of each day rather than a fixed clock-based cutoff. This approach was adopted to account for nocturnal snacking. For example, a last meal at 9 PM and a first meal at 8 AM the following day yields an NFI of 11 hours. If one additionally had a snack at 1 AM the longest fasting interval will shift to 1 AM to 8 AM, resulting in an NFI of 7 hours. A definition of ≥50 kcal was used to represent an eating episode (Gibney and Wolever, 1997). NFIs were averaged across the two dietary recall days and were categorized into three groups at each timepoint (i.e., short: < 25^th^ percentile, middle: 25^th –^75^th^ percentile, and long: > 75^th^ percentile).

#### Chronotype indicators

Several indicators have been proposed for chronotype, including dim-light melatonin onset (DLMO), sleep midpoint (i.e., the mid-point of the sleep interval), and breakfast time. Of these, DLMO is considered the gold standard, and sleep midpoint is a robust alternative. We used self-reported breakfast time as a proxy for chronotype, as sleep midpoint was not available, and this variable was categorized by median split to distinguish eveningness from morningness. In a sensitivity analysis, we instead used sleep midpoint for chronotype.

Breakfast time, while generally less preferred, is a practical proxy for chronotype: It requires neither costly laboratory testing nor sleep information, yet shows a moderate correlation with chronotype (r = -0.469 for free days; r = -0.325 for work days) (Bazzani *et al*., 2022).

#### Depressive symptoms

Depressive symptoms were assessed using the Center for Epidemiologic Studies-Depression Scale (CES-D), a 20-item self-report measure of depressive symptoms that are rated on a Likert scale ranging from 0 to 3, yielding a maximum score of 60 (i.e., higher scores indicate greater severity of depressive symptoms). The total score was treated as a continuous variable after log-transformation, and geometric mean ratios (GMRs) were presented to quantify the association between NFIs and CES-D scores. A cutoff of 16 was additionally applied to classify clinically significant depression in a sensitivity analysis (Radloff, 1977; Tandon *et al*., 2012).

#### Pittsburgh Sleep Quality Index

The Pittsburgh Sleep Quality Index (PSQI) is a self-rated questionnaire that assesses sleep quality and disturbances (Buysse *et al*., 1989). PSQI has 7 components with 0-3 scores, yielding a global score ranging from 0 to 21, with higher values indicating poorer sleep quality.

#### Sociodemographic and health-related characteristics

Baseline information included maternal age, racial and ethnic identities, pre-pregnancy BMI, parity, education, household income, marital status, insurance, and employment. A composite socioeconomic status (SES) variable was calculated by averaging household income (z-scored) and educational level (z-scored; categorized by 1: grade school, 2: high school, 3: college, 4: 4-year college, and 5: graduate degree), and we re-standardized the resulting value to yield a distribution with a mean of 0 and standard deviation of 1 (Dougherty *et al*., 2020).

### Statistical analysis

First, a mixed effects linear model was applied with random intercepts for individuals to estimate GMRs for the longitudinal associations between NFIs and depressive symptoms, represented by log-transformed CES-D scores. An interaction term for NFIs and timepoints (i.e., early-pregnancy, mid-pregnancy, and 6-months postpartum) was included in this model., which further adjusted for the following covariates: Breakfast time (a time-varying measure of chronotype: dichotomized by median split), maternal age (continuous), marital status (binary: married and unmarried), parity (binary: nulliparous or multiparous), pre-pregnancy BMI (binary: <30 and ≥30), employment (binary: working or not working) and SES (continuous). Estimated marginal means were calculated by averaging individual predicted probabilities across all observations.

Second, we dichotomized CES-D (≥16 or <16) and used this dichotomized indicator in a mixed effect Poisson regression model to obtain prevalence ratios with random intercepts for individuals. The same covaries were included, and estimated marginal probabilities were calculated using the same approach.

### Sensitivity analyses

Using a subgroup of the sample with sleep information from the PSQI, we ran the same mixed effects linear model described above to evaluate the association between NFIs and depressive symptoms, but this time covarying for sleep quality (PSQI total score: continuous) and sleep midpoint (a measure of chronotype: dichotomized by median split) rather than breakfast time. This analysis was restricted to participants who had the PSQI, NFIs, and CES-D at baseline and at least one other timepoint. The maternal demographics and a comparison between the sensitivity and main analysis were visualized. Participants reporting wake times after 12:00 PM were excluded because they were likely to be night-shift workers (Wirth *et al*., 2021). Estimated marginal means were also calculated.

All analyses were conducted using R version 4.5.0. Significance level was set as 0.05.

## Results

### Participants

The study flow diagram is presented in Figure 1, and maternal baseline characteristics are summarized in Table 1. Participants with longer NFIs were younger and had a higher prevalence of obesity (BMI ≥30) compared to the short and middle NFI group. Higher SES was observed among participants with middle NFIs. Information on NFIs, breakfast time, and CES-D scores is summarized in Table 2. NFIs remained largely stable with a slight increase (12.0 hours at early-pregnancy to 13.0 hours at postpartum) throughout the study period, whereas breakfast time remained stable. CES-D scores declined over time (66 scores at early-pregnancy to 44 scores at postpartum).

**Fig. 1.**
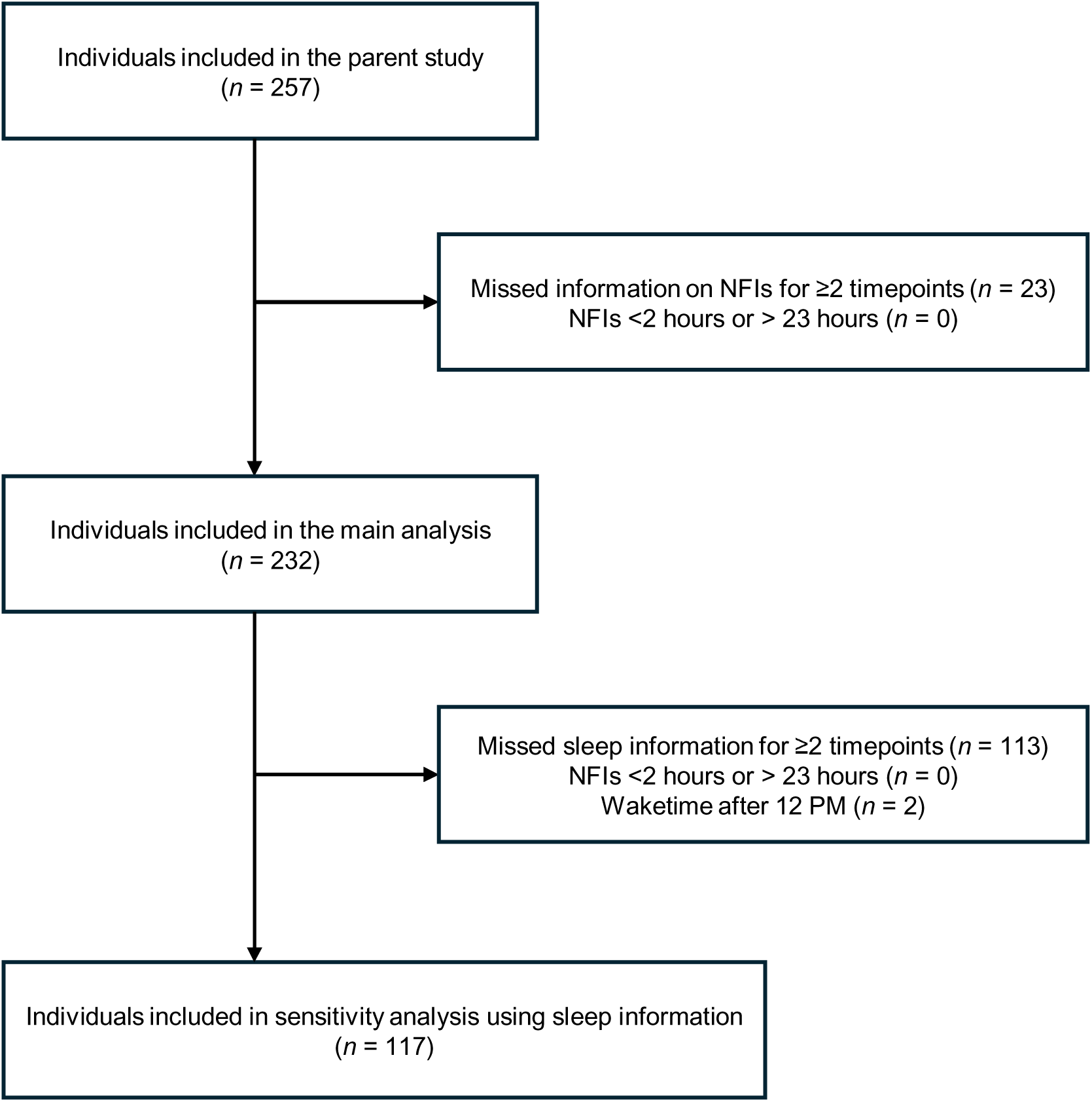
The diagram shows the study flow

**Table. 1.** Maternal demographics across nighttime fasting interval length categories.

|  |  | Short<br>*n* = 58<sup>a</sup> | Middle<br>*n* = 123<sup>a</sup> | Long<br>*n* = 51<sup>a</sup> | *p*-value<sup>b</sup> |
| --- | --- | --- | --- | --- | --- |
| **Age (years)** | Mean (*SD*) | 29 (5.9) | 29 (5.4) | 26 (4.8) | 0.001 |
| **Race/Ethnicity** | *n* (%) |  |  |  | 0.4 |
| Non-Hispanic Black |  | 25 (43%) | 54 (44%) | 27 (53%) |  |
| Non-Hispanic White |  | 28 (48%) | 65 (53%) | 23 (45%) |  |
| Others |  | 5 (9%) | 4 (3%) | 1 (2%) |  |

|  |  | **Short**<br>*n* = 58<sup>a</sup> | **Middle**<br>*n* = 123<sup>a</sup> | **Long**<br>*n* = 51<sup>a</sup> | ***p*-value<sup>b</sup>** |
| --- | --- | --- | --- | --- | --- |
| **BMI** | *n* (%) |  |  |  | 0.045 |
| <30 |  | 27 (47%) | 58 (47%) | 14 (27%) |  |
| ≥30 |  | 31 (53%) | 65 (53%) | 37 (73%) |  |
| **Parity** | *n* (%) |  |  |  | 0.2 |
| Nulliparous |  | 16 (28%) | 51 (41%) | 17 (33%) |  |
| Multiparous |  | 42 (72%) | 72 (59%) | 34 (67%) |  |
| **Socioeconomic status** | Mean (*SD*) | -0.22 (0.73) | 0.14 (0.91) | -0.16 (1.33) | 0.004 |
| **Education** | *n* (%) |  |  |  |  |
| High school or less |  | 21 (36%) | 30 (24%) | 22 (43%) |  |
| Some college |  | 33 (57%) | 66 (54%) | 25 (49%) |  |
| Graduate school |  | 4 (7%) | 27 (22%) | 4 (8%) |  |
| **Household income** | *n* (%) |  |  |  |  |
| Less than \$30,000 | | 41 (71%) | 70 (57%) | 39 (78%) | |
| \$30,001 to \$50,000 | | 7 (12%) | 8 (7%) | 4 (8%) | |
| \$50,001 to \$70,000 | | 5 (9%) | 8 (7%) | 4 (8%) | |
| \$70,001 to \$100,000 | | 1 (2%) | 20 (16%) | 3 (6%) | |
| Greater than \$100,000 | | 4 (7%) | 17 (14%) | 0 (0%) | |
| **Marital status** | *n* (%) | 17 (29.3%) | 51 (41.5%) | 11 (21.6%) | 0.028 |
| **Insurance** | *n* (%) |  |  |  | 0.022 |
| Provided by Employer |  | 15 (26%) | 56 (46%) | 13 (25%) |  |
| Medicare/Medicaid |  | 39 (67%) | 59 (48%) | 37 (73%) |  |
| Personal Policy |  | 2 (3%) | 3 (2%) | 1 (2%) |  |
| No Health Insurance |  | 2 (3%) | 5 (4%) | 0 (0%) |  |
| **Employment** | *n* (%) |  |  |  | 0.022 |
| Not working |  | 28 (48%) | 38 (31%) | 25 (49%) |  |
| Working |  | 30 (52%) | 85 (69%) | 26 (51%) |  |

Continuous variables are presented as mean (SD) or median (IQR) based on distribution; categorical variables are presented as *n* (%). <sup>a</sup> Mean (SD); *n* (%), <sup>b</sup> Kruskal-Wallis rank sum test; Fisher's exact test; Pearson's Chi-squared test.

**Table. 2.** Descriptive statistics for variables of interest across timepoints.

|  |  | early-pregnancy<br>*n* = 232 | mid-pregnancy<br>*n* = 218 | postpartum<br>*n* = 202 |
| --- | --- | --- | --- | --- |
| **Nighttime fasting interval (hours)** | Median (IQR) | 12.0 (10.6, 14.0) | 12.5 (11.0, 14.5) | 13.0 (11.0, 15.0) |
| **Breakfast time (hour)** | Median (IQR) | 9:18 (8:18, 10:30) | 9:18 (8:18, 10:18) | 9:30 (8:30, 11:00) |
| **CES-D<sup>a</sup> score** | Median (IQR) | 10.0 (5.0, 17.5) | 9.0 (4.0, 16.0) | 8.0 (4.0, 14.0) |
| **Depression (CES-D<sup>a</sup> ≥16)** | *n* (%) | 66 (28.4%) | 60 (27.5%) | 44 (21.8%) |

<sup>a</sup> CES-D: Center for Epidemiologic Studies Depression Scale

### Main results

The adjusted model results are presented in Table 3. NFIs were not associated with depressive symptoms during pregnancy or the postpartum period. Also, the interaction between NFIs and each timepoint was not statistically significant (*p* = 0.87). The estimated marginal means of CES-D scores are shown in Figure 2. Although the associations were not significant, the point estimates suggested long NFIs showed lower estimated CES-D scores relative to short or middle NFIs at mid-pregnancy, whereas the associations did not show consistent directions in early-pregnancy and postpartum.

**Fig. 2.**
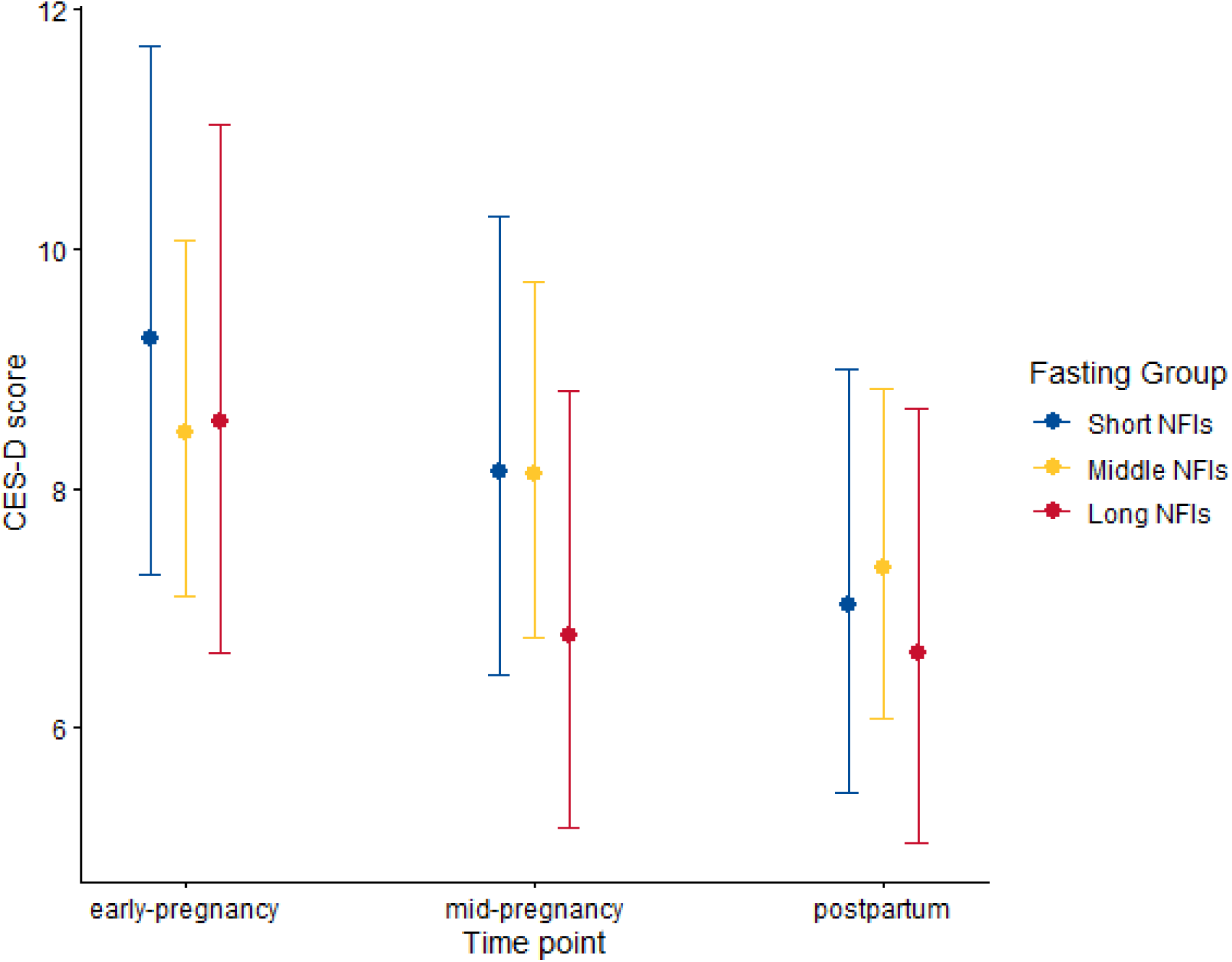
Estimated marginal means of CES-D scores across the study period, calculated from the adjusted model. Blue represents short NFI, yellow represents middle NFI, and red represents long NFI. CES-D: Center for Epidemiologic Studies Depression Scale; NFI: nighttime fasting interval

**Table. 3.** Results from adjusted linear model testing the association between nighttime fasting intervals and depressive symptoms (CES-D total score)

|  | Short interval |  | Middle interval (ref) |  | Long interval |  |
| --- | --- | --- | --- | --- | --- | --- |
| Time point | Geometric mean (*n*) | GMR (95% CI); *p*<sup>a</sup> | Geometric mean (*n*) | GMR | Geometric mean (*n*) | GMR (95% CI); *p*<sup>a</sup> |
| early-pregnancy | 9.6 (n=58) | 1.09 (0.85, 1.40);<br>*p*=0.514 | 8.0 (n=123) | 1.0 | 8.3 (n=51) | 1.01 (0.77, 1.32);<br>*p*=0.936 |
| mid- | 8.1 (n=59) | 1.00 (0.78, 1.29); | 7.1 (n=113) | 1.0 | 7.8 (n=46) | 0.84 (0.64, 1.11); |

|  | Short<br>interval |  | Middle<br>interval<br>(ref) |  | Long<br>interval |  |
| --- | --- | --- | --- | --- | --- | --- |
| Time point | Geometric<br>mean (n) | GMR (95% CI);<br>*p*<sup>a</sup> | Geometric<br>mean (n) | GMR | Geometric<br>mean (n) | GMR (95% CI); *p*<sup>a</sup> |
| pregnancy |  | p=0.978 |  |  |  | p=0.226 |
| postpartum | 6.7 (n=52) | 0.96 (0.74, 1.25);<br>p=0.758 | 6.8 (n=104) | 1.0 | 7.1 (n=46) | 0.91 (0.69, 1.21);<br>p=0.512 |

<sup>a</sup> GMR: Geometric Mean Ratio; CI: Confidence Interval

NFIs were not significantly associated with the probability of depression (Table 4). The estimated marginal probabilities of depression are presented in Figure 3. Although the differences were not statistically significant, short NFIs were associated with the highest probability of depression, approximately 10 percents higher than the long NFIs, during mid-pregnancy (estimated marginal probability for short NFIs: 0.33 (95% CI 0.23–0.47) vs. long NFIs: 0.20 (95% CI 0.11–0.33)).

**Fig. 3.**
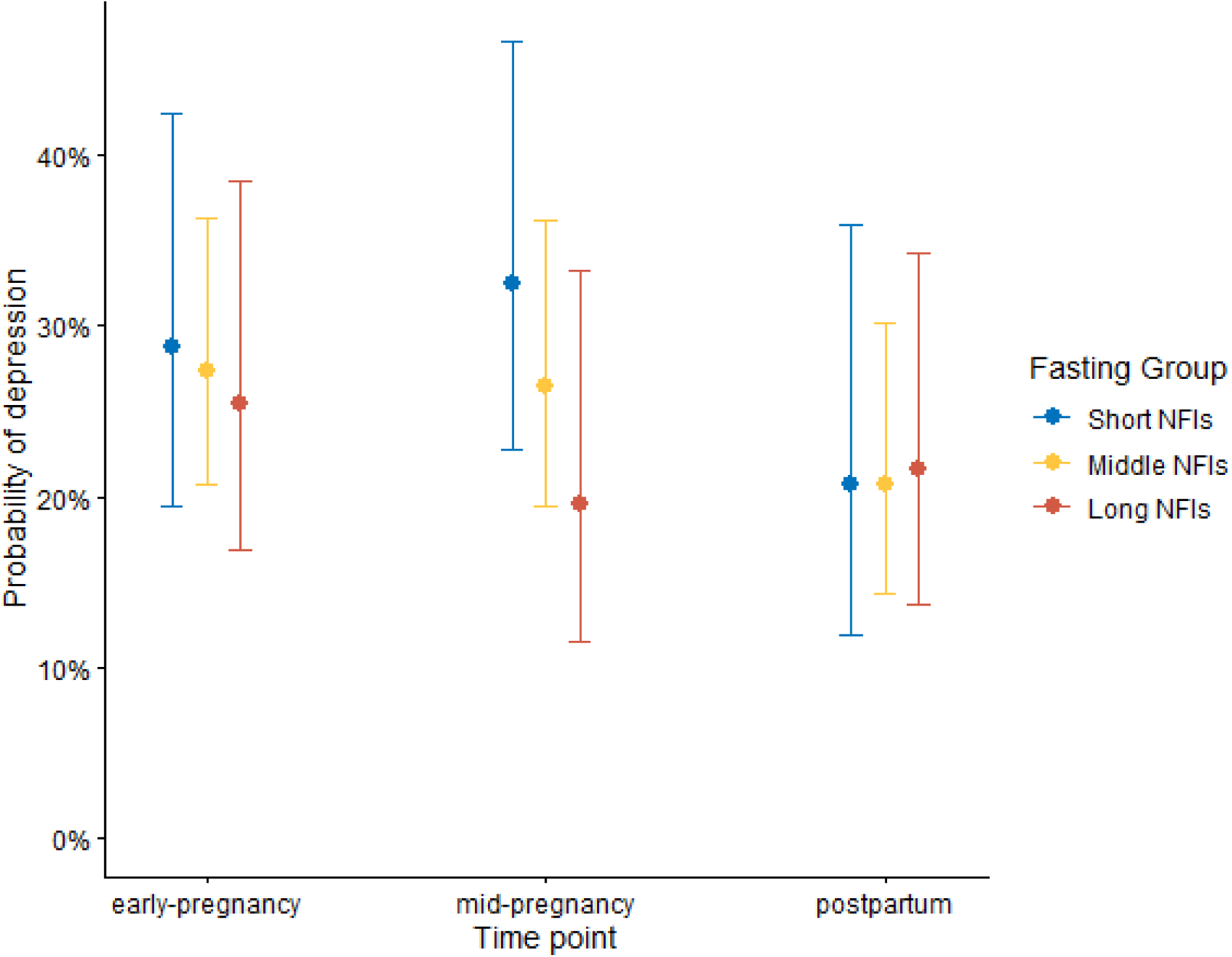
Estimated marginal probabilities of depression across the study period, calculated from the adjusted model. Blue represents short NFI, yellow represents middle NFI, and red represents long NFI. NFI: nighttime fasting interval

**Table. 4.** Results from adjusted Poisson model testing the association between nighttime fasting intervals and.

|  | Short<br>interval |  | Middle<br>interval<br>(ref) |  | Long<br>interval |  |
| --- | --- | --- | --- | --- | --- | --- |
| Time point | Prevalence | PR (95% CI); *p*<sup>a</sup> | Prevalence | PR | Prevalence | PR (95% CI); *p*<sup>a</sup> |
| early-pregnancy | 17/58 | 1.01 (0.89, 1.16);<br>p=0.837 | 33/123 | 1.0 | 16/51 | 0.98 (0.86, 1.12);<br>p=0.760 |
| mid-pregnancy | 20/59 | 1.06 (0.93, 1.22);<br>p=0.368 | 28/113 | 1.0 | 12/46 | 0.93 (0.82, 1.06);<br>p=0.271 |
| postpartum | 10/52 | 1.00 (0.87, 1.15);<br>p=0.991 | 22/104 | 1.0 | 12/46 | 1.01 (0.89, 1.14);<br>p=0.891 |

|  | Short interval |  | Middle interval (ref) |  | Long interval |  |
| --- | --- | --- | --- | --- | --- | --- |
| Time point | Prevalence | PR (95% CI); *p*<sup>a</sup> | Prevalence | PR | Prevalence | PR (95% CI); *p*<sup>a</sup> |

<sup>a</sup> PR: Prevalence Ratio; CI: Confidence Interval

### Sensitivity analysis

Sensitivity analysis included participants with sleep information (*n* = 117; Figure 1). Maternal baseline demographics and a comparison of baseline demographics are shown in Supplementary tables 1 and 2, respectively. A comparison table showed participant characteristics were similar between the main and subsample with sleep quality data (Supplementary table 2). Mean sleep quality remained relatively stable (PSQI: *M* [*SD*] = 6.53 [3.56] scores), whereas the sleep midpoint shifted earlier across the perinatal period (Supplementary table 3; 3:18 AM at early-pregnancy, 3:00 AM at mid-pregnancy, 2:36 AM at postpartum). Long NFIs in mid-pregnancy showed a lower GMR compared to middle NFIs (Table 5). Although differences at other timepoints were minimal, the estimated CES-D score in the long NFI group was approximately 4 points lower than that in the middle NFI group during mid-pregnancy (Figure 4; estimated marginal mean for middle NFIs: 10.9 (95% CI 8.11–14.4); long NFIs: 6.4 (95% CI 4.3–9.4)).

**Fig. 4.**
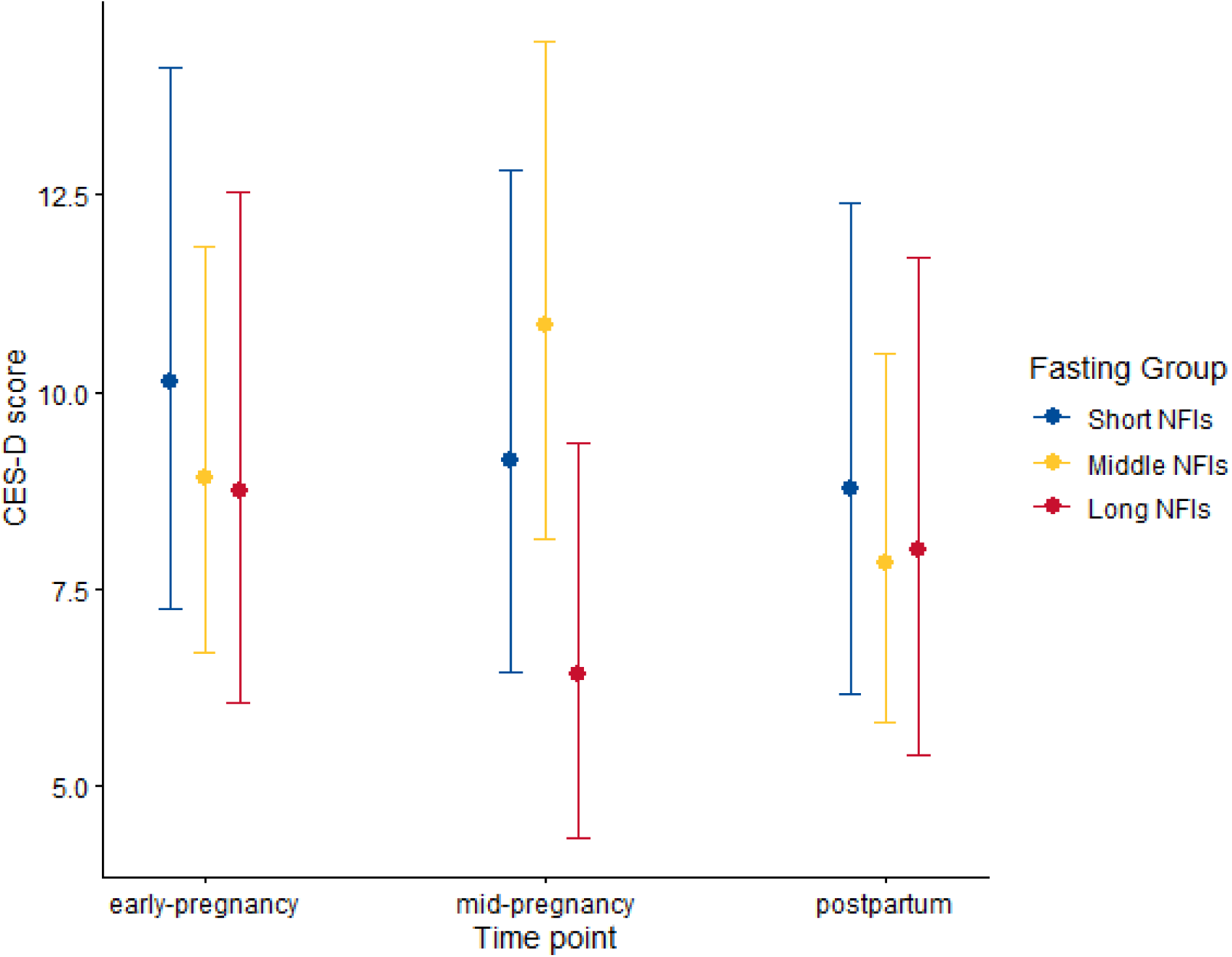
Estimated marginal means of CES-D scores across the study period, calculated from the adjusted model including sleep information. Blue represents short NFI, yellow represents middle NFI, and red represents long NFI. CES-D: Center for Epidemiologic Studies Depression Scale; NFI: nighttime fasting interval

**Table. 5.** Results from adjusted linear model examining the association between NFIs and depressive symptoms (CES-D total score) among those with sleep information.

|  | Short interval |  | Middle interval (ref) |  | Long interval |  |
| --- | --- | --- | --- | --- | --- | --- |
| Time point | Geometric mean (*n*) | GMR (95% CI); *p*<sup>a</sup> | Geometric mean (*n*) | GMR | Geometric mean (*n*) | GMR (95% CI); *p*<sup>a</sup> |
| early-pregnancy | 8.6 (n=31) | 1.13 (0.82, 1.55);<br>*p*=0.469 | 8.5 (n=58) | 1.0 | 8.3 (n=28) | 0.99 (0.71, 1.38);<br>*p*=0.960 |
| mid-pregnancy | 9.4 (n=29) | 0.83 (0.60, 1.15);<br>*p*=0.268 | 8.1 (n=56) | 1.0 | 7.5 (n=24) | 0.59 (0.41, 0.84);<br>*p*=0.004 |
| postpartum | 8.0 (n=29) | 1.09 (0.78, 1.52);<br>*p*=0.625 | 6.1 (n=48) | 1.0 | 6.8 (n=21) | 1.02 (0.70, 1.48);<br>*p*=0.912 |

<sup>a</sup> GMR: Geometric Mean Ratio; CI: Confidence Interval

## Discussion and conclusions

In this secondary analysis, NFIs were not significantly associated with depressive symptoms during pregnancy or postpartum. However, estimated marginal probabilities suggested that participants with short NFIs relative to long NFIs had an approximately 10 percents higher probability of depression during mid-pregnancy. In a sensitivity analysis restricted to participants with sleep information, longer NFIs were significantly associated with lower depressive symptoms compared to middle NFIs during mid-pregnancy.

The Minimal Clinically Important Difference (MCID) for CES-D is generally considered to be approximately 6.6 points (Haase, Winkeler and Imgart, 2022). While sensitivity analyses yielded statistically significant findings, the estimated between-group differences did not reach this threshold, warranting caution in drawing firm conclusions. Nevertheless, mid-pregnancy may represent a particularly susceptible period where NFIs could affect maternal mental health, compared to other perinatal timepoints.

The unique physiological changes may help explain the present findings. For example, the maternal gut microbiome undergoes substantial remodeling from the first to the third trimester (Ali and Kunugi, 2020). Early-pregnancy microbiota resembles that of non-pregnant adults, characterized by high diversity and stability. However, by the third trimester, individuals experience a profound reduction in intestinal microbiome diversity, and colonization patterns resemble those of obesity, leading to a heightened inflammatory condition and increased insulin resistance (Ali and Kunugi, 2020; Codoñer-Franch *et al*., 2023; Hu *et al*., 2026). In addition, the gut microbiota and circadian rhythm are closely linked. Circadian disruption leads to proinflammatory taxa and a decreased abundance in microbiota-mediated pathways crucial for brain functioning, including tryptophan biosynthesis for serotonin production (Codoñer-Franch *et al*., 2023). Inflammation also plays a key role throughout the perinatal period. The first trimester is characterized by a pro-inflammatory state, followed by an anti-inflammatory state in the second trimester, and a return to a pro-inflammatory state in the third trimester (Mor and Cardenas, 2010; Hu *et al*., 2026). These changes are driven by implantation, placentation, and preparation for delivery. Following childbirth, both gut microbiome composition and inflammatory states gradually recover over the course of several months postpartum (Bränn *et al*., 2022; Weerasuriya *et al*., 2023). Therefore, the influence of these physiological changes is expected to be minimal at 6 months postpartum in the present study.

In this regard, short NFIs are often accompanied by night eating, which disrupts circadian rhythm and gut microbiome composition, collectively accelerating inflammation and contributing to depressive symptoms. Conversely, long NFIs have been shown to reduce inflammation by inhibiting the translocation of bacterial toxins from the gut (Ali and Kunugi, 2020). Long NFIs also generate sharp feeding-fasting cycles that consolidate circadian activation of various metabolic pathways (Loy *et al*., 2017), and further exert neuroprotective effects by upregulating neurotrophic factor expression and reducing proinflammatory cytokine levels (Codoñer-Franch *et al*., 2023). The higher variability in their associations with depressive symptoms during mid-pregnancy may be attributable to the dynamic interplay of gut microbiome composition, circadian rhythm, and inflammatory states characteristic of this period. This complexity throughout pregnancy may explain why the present study did not show the U-shaped association between NFIs and depression found in non-pregnant populations (Chen *et al*., 2025).

This study highlights several important directions for future research. First, the use of objective measures, such as DLMO, would reduce residual confounding attributable to chronotype. Second, the present findings suggest that the effect of NFIs on depressive symptoms may be more pronounced during late-pregnancy. Future studies should therefore examine this period as a potentially critical window for intervention.

### Strengths and limitations

Several strengths of this study should be noted. First, this prospective cohort study included perinatal populations, which have not been well studied in previous research. The longitudinal design with repeated measures also allowed us to examine the association between NFIs and perinatal depression over time. The present study also adjusted for breakfast time in the main analysis and sleep midpoint in the sensitivity analysis as a practical proxy for chronotype.

Limitations include the small sample size relative to recent studies that leveraged data from nationwide surveys or large cohorts (Loy *et al*., 2017; Zhang *et al*., 2024; Chen *et al*., 2025). Additionally, the use of self-reported measures for fasting intervals, breakfast time, sleep, and depression may have introduced recall bias and shared method variance. Moreover, we did not account for diet quality, which could have resulted in residual confounding (Baskin *et al*., 2015). Finally, as this secondary analysis was restricted to individuals with overweight or obesity, findings may not generalize to broader populations.

### Conclusions

In this overweight and obese perinatal individuals, NFIs were not significantly associated with depressive symptoms. However, we observed high variability between short and long NFIs in mid-pregnancy. Future studies should incorporate objective measures for sleep or other circadian indicators (e.g., DLMO) and more frequent assessments (e.g., third trimester).

## Data Availability

All data produced in the present study are available upon reasonable request to the authors

## Declarations

### Ethics approval

The University of Pittsburgh’s Institutional Review Board approved all study procedures (PRO11070083).

### Consent to participate

Informed consent was obtained from all individual participants included in the study. Data availability statement

### Data availability

The de-identified individual participant database, as well as a data dictionary and relevant related documents (e.g., study protocol) are available with reasonable request. Requests can be made with the corresponding author.

### Author contributions

Conceptualization: Takashi Yoshimasu, Michele D. Levine, Marquis Hawkins; Data curation and Investigation: Riley J. Jouppi, Christine C. Call, Lydia B. Brown, Rachel P. Kolko Conlon, Michele D. Levine, Marquis Hawkins; Formal analysis: Takashi Yoshimasu, Marquis Hawkins; Funding acquisition: Michele D. Levine, Marquis Hawkins; Methodology: Takashi Yoshimasu, Christine C. Call, Michele D. Levine, Marquis Hawkins; Project administration: Michele D. Levine, Marquis Hawkins; Supervision: Michele D. Levine, Marquis Hawkins; Validation: Marquis Hawkins; Visualization: Takashi Yoshimasu, Marquis Hawkins; Writing original draft: Takashi Yoshimasu; Writing review & editing: Takashi Yoshimasu, Christina M. Personette, Namhyun Kim, Rya Clifton, David Phan, Mehak Behal, Riley J. Jouppi, Andrea B. Goldschmidt, Christine C. Call, Lydia B. Brown, Shruti Kinkel-Ram, Rachel P. Kolko Conlon, Michele D. Levine, Marquis Hawkins

## Acknowledgements

Data collection was supported by R01 HL132578 (PI: Michelle D. Levine). Dr. Takashi is supported by Graduate Scholarship for Degree Seeking Students by the Japan Student Services Organization (JASSO) Student Exchange Support Program and Global Grant Scholarship by the Rotary Foundation (GG2579376). Dr. Call is supported by The National Institute of Diabetes and Digestive and Kidney Diseases (K23 DK136927). Dr. Kinkel-Ram is supported by funding from the National Heart Lung and Blood Institute (5T32HL007560).

